# Association between oral health and hyperuricemia in Korean adults: Korea National Health and Nutrition Examination Survey 2016-2019

**DOI:** 10.1101/2022.01.18.22269502

**Authors:** Junyong Park, Minkook son, Sung Won Lee, Won Tae Chung, Sang Yeob Lee

## Abstract

Hyperuricemia plays an essential role in the development of gout. Despite the clinical importance of hyperuricemia, a direct relationship between oral health and hyperuricemia has not been established. Therefore, we aim to investigate the association between oral health and hyperuricemia in Korean adults.

We selected 17,584 subjects from the Korea National Health and Nutrition Examination Survey 2016–2019 database. Oral health-related variables included the number of dental caries, regular tooth brushing, use of secondary oral products, and regular dental examination. The odds ratio (OR) and 95% confidence intervals (CIs) for hyperuricemia were calculated using a multivariable-adjusted logistic regression model.

In all subjects, oral health status with dental caries and oral health behaviors, including tooth brushing; secondary oral products; and regular dental examination, were significantly associated with hyperuricemia. The adjusted OR and 95% CIs for hyperuricemia comparing more than three dental caries with no dental caries were 1.28 (1.08–1.51). The adjusted OR and 95% CIs for hyperuricemia in regular tooth brushing, use of secondary oral products, and regular dental examination were 0.78 (0.67–0.91), 0.91 (0.82–1.00), and 0.86 (0.78–0.95), respectively. Notably, the association between oral health and hyperuricemia was more prominent in male subjects. In addition, when subjects were grouped by the scoring system regarding oral health, the prevalence of hyperuricemia was lower in groups with a better oral health score.

In conclusion, we demonstrated that oral health status and behaviors were associated with hyperuricemia, especially in males. Further complementary studies are needed to confirm the definite association between oral health and hyperuricemia.

## Introduction

Hyperuricemia is a metabolic disease in which uric acid level remains excessively high. Hyperuricemia plays an essential role in developing gout and is well known as a prerequisite state of gout. In addition, hyperuricemia has important clinical implications because it is considered a risk factor for coronary heart disease, hypertension, insulin resistance, stroke, and death [1–4]. Nevertheless, the prevalence rate of hyperuricemia is 11.4% in Korea and 12.7% in the United states [5,6] and the prevalence rate of gout is increasing rapidly in Korea [7]

Oral cavity is a complex environment with various microorganisms [8]. Unless it is managed appropriately, inflammatory conditions, such as dental caries and periodontitis, may occur. Furthermore, oral health is known to be related to pneumonia and cardiovascular diseases as well as oral diseases [9,10]. Oral health is critically affected by oral hygiene and can be improved by regular self-management. Therefore, it would be important to investigate specific diseases related to oral health and prevent them [11].

Despite the clinical importance of hyperuricemia, a direct relationship between oral health and hyperuricemia has not been established. Therefore, we aim to investigate the association between oral health and hyperuricemia in Korean adults using the nationwide population-based Korea National Health And Nutrition Examination Survey (KNHANES) database.

## Material and methods

### Data source and study population

The KNHANES is a cross-sectional survey and a nationally representative database of the Korean population managed by the Korea Centers for Disease Control and Prevention (KCDC) [12]. The KNHANES database includes physical examinations, blood test results, and health-related interviews including oral health behaviors. To conduct the KNHANES, the trained staff interview subjects and apply the standardized health examination protocols [13]. This study used data from the KNHANES 2016–2019 because this database contains the blood test results for uric acid level. Informed consent was obtained from all participants and the KNHANES was approved by the Institutional Review Board of the KCDC. This study used the anonymized KNHANES data, and the protocol of this study was approved by the Institutional Review Board of Dong-A university hospital (DAUHIRB-EXP-21-106).

From 23,335 subjects in the KNHANES 2016–2019 database, we excluded 3,255 subjects without uric acid levels. The subjects under the age 20 (n = 2,063), with a pregnancy (n = 69), and with missing values (n = 364) were also excluded. Finally, this study included 17,584 subjects (7,831 males and 9,753 females). Subsequently, we investigated the association between oral health and uric acid. A flow of this study population is described in Fig 1.

**Fig 1.**
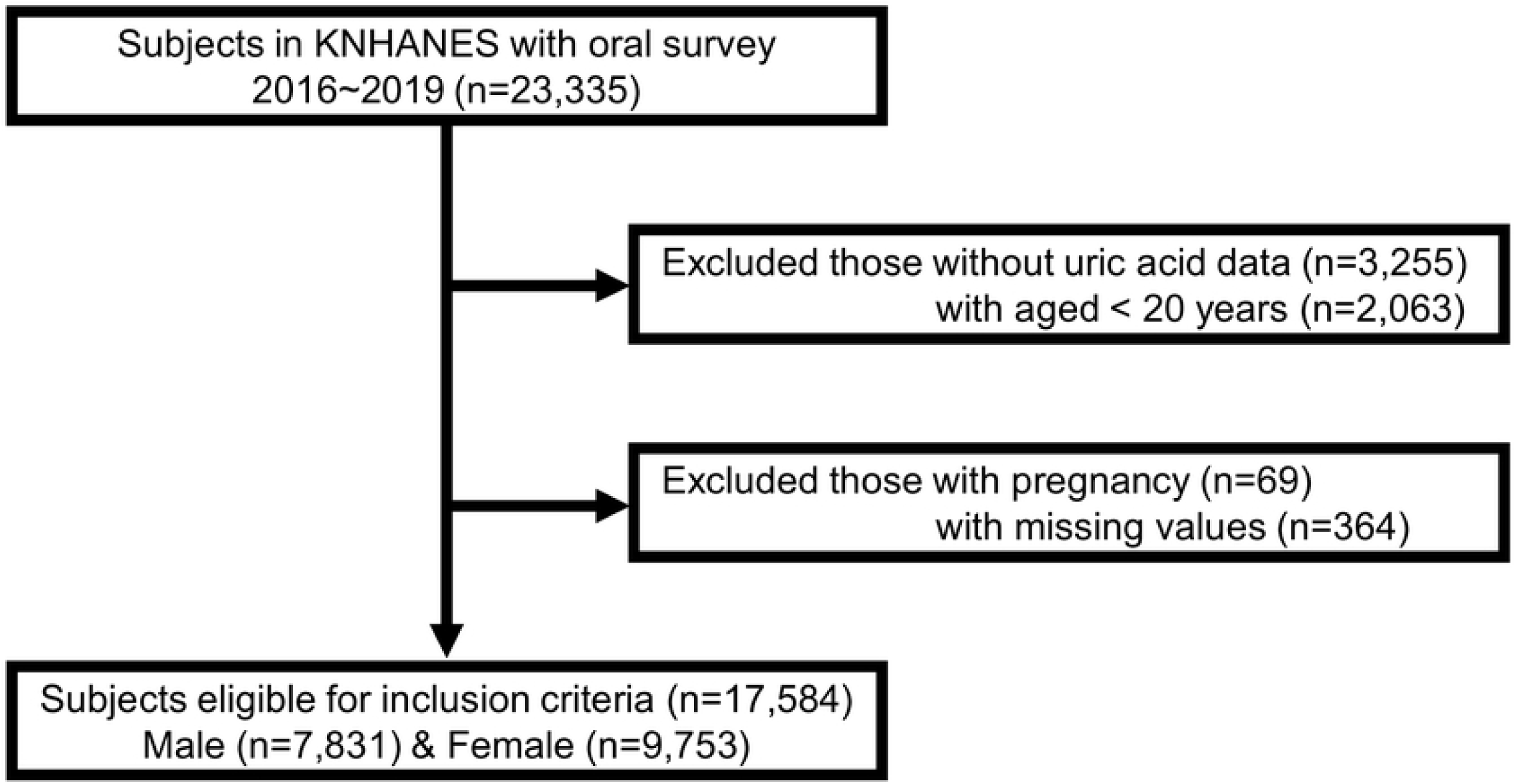
Flow of the study population.

### Oral health-related variables’ definition

The oral health examination program was composed of oral examinations conducted by trained dentists and self-reported questionnaires. The number of dental caries was categorized as 0, 1–2, and ≥3. Oral health-related questionnaires included the questions on the time of day when subjects brushed their teeth and used secondary oral products as well as the presence of regular dental examination as oral health behaviors. The time of day for tooth brushing was classified as before or following breakfast, lunch, and dinner or before bedtime and after snack. We calculated the frequency of daily toothbrushing a day. The number of tooth brushing was categorized as ≤1, 2, and ≥3. Secondary oral products included the following: dental floss, mouthwash, interdental brushes, electric toothbrushes, irrigation devices, tongue cleaners, end-tufted brushes, and special devices for dentures. The presence of regular dental examination within a year was also evaluated.

To evaluate the association between oral health and hyperuricemia, we calculated the oral health score by scoring the oral health-related variables and summing them altogether. A schematic diagram for the oral health score is presented in S1 Table. For example, the higher oral health score suggests better oral hygiene, and the lower oral health score indicates poor oral hygiene.

### Clinical variables measurement and definition

Body mass index (BMI) was calculated using the formula weight (kg) divided by height squared (m^2^). All subjects fasted for at least eight hours before blood sampling. Blood samples were processed and refrigerated immediately for transportation to the Central Testing Institute (NeoDin Medical Institute, Seoul, Korea). Uric acid, fasting glucose, total cholesterol, triglycerides, high-density lipoprotein cholesterol (HDL-C), and creatinine were measured using the Hitachi Automatic Analyzer 7600-210 (Hitachi, Japan). Estimated glomerular filtration rate (eGFR) was calculated using the modification of diet in renal disease equation [14]. Hyperuricemia was defined as serum uric acid level ≥ 7.0 mg/dL in male and ≥ 6.0 mg/dL in female [15].

Hypertensive status was categorized into three groups: (a) hypertension was defined as systolic blood pressure (SBP) ≥ 140 mmHg, diastolic blood pressure (DBP) ≥ 90 mmHg, or taking anti-hypertensive medication; (b) pre-hypertension was defined as 120 mmHg ≤ SBP <140 mmHg or 80 mmHg ≤ DBP < 90 mmHg; and (c) normal was defined as SBP < 120 mmHg and DBP < 80 mmHg [16]. Diabetic status was categorized into three groups: (a) diabetes was defined as fasting blood sugar (FBS) ≥ 126 mg/dL or taking anti-diabetic medication; (b) pre-diabetes was defined as 100 mg/dL ≤ FBS < 126 mg/dL; and (c) normal was defined as FBS < 100 mg/dL [17]. Using the dyslipidemia criteria for Koreans, Dyslipidemia status was categorized into two groups with cut-off values as follows: total cholesterol ≥ 240 mg/dL, triglycerides ≥ 200 mg/dL, and HDL-C ≤ 40 mg/dL [18].

Income level was categorized into four groups. Smoking status was categorized into current smokers, ex-smokers, and non-smokers. Alcohol consumption status was categorized into two groups: (a) non-consumers, with no alcohol consumption in the past year or less than once a month, and (b) consumers, with alcohol consumption more than once a month. Physical exercise status was categorized into two groups: (a) the regular exercise group, who performed moderately hard exercise at least 150 minutes a week, hard exercise at least 75 minutes a week, or mixed exercise equivalent to the above level (1 minute of hard exercise equivalent to 2 minutes of moderately hard exercise) and (b) non-regular exercise group, who performed physical activity less than the above-mentioned level.

### Statistical analysis

The characteristics of subjects were analyzed according to their uric acid levels. Continuous variables are presented as the mean and standard deviations, and categorical variables are presented as the number of cases with percentage. The logistic regression analysis was performed and odds ratio (OR) with 95% confidence intervals (CIs) for hyperuricemia were calculated to investigate the association between oral health and hyperuricemia. The multivariable-adjusted logistic regression analysis was adjusted for age, sex, income level, BMI, GFR, hypertension, diabetes, dyslipidemia status, smoking status, alcohol consumption, and regular exercise status. Data processing and statistical analyses were performed using R version 4.0.5. (https://www.r-project.org/) and SPSS software version 20 (IBM, SPSS Inc., NY, USA). The P-value < 0.05 was considered statistically significant.

## Results

### Baseline characteristics of the study population

Baseline characteristics of the subjects according to uric acid levels are presented in Table 1. There were significant differences between no hyperuricemia and hyperuricemia groups except in the exercise variable. Mean values of uric acid levels in each group were 4.7 and 7.5, respectively. Hyperuricemia group was identified in young and male. In addition, compared to no hyperuricemia group, hyperuricemia group had higher BMI, SBP, DBP, fasting blood glucose, total cholesterol, and triglyceride levels but lower HDL-C and GFR levels. Comorbidities, including hypertension; diabetes; and dyslipidemia, were more common in the hyperuricemia group. The higher number of dental caries and the low number of tooth brushing were more common in the hyperuricemia group. The use of secondary oral products and regular dental examination were more frequent in the no hyperuricemia group. Baseline characteristics of subjects divided into male and female are presented in S2 and S3 Tables.

**Table 1.** Baseline characteristics of subjects according to uric acid level.

| Total<br>(n = 17,584) | No hyperuricemia<br>(n =15,219) | Hyperuricemia<br>(n = 2,365) | P-value |
| --- | --- | --- | --- |
| Age (years) | 51.9 ± 16.3 | 50.0 ± 17.8 | < 0.001 |
| Sex |  |  | < 0.001 |
| Male | 6218 (40.9%) | 1613 (68.2%) |  |
| Female | 9001 (59.1%) | 752 (31.8%) |  |
| Income (quartile) |  |  | 0.03 |
| 1<sup>st</sup> | 3747 (24.6%) | 625 (26.4%) |  |
| 2<sup>nd</sup> | 3879 (25.5%) | 564 (23.8%) |  |
| 3<sup>rd</sup> | 3808 (25.0%) | 627 (26.5%) |  |
| 4<sup>th</sup> | 3785 (24.9%) | 549 (23.2%) |  |
| BMI (kg/m<sup>2</sup>) | 23.7 ± 3.4 | 25.8 ± 3.8 | < 0.001 |
| SBP (mmHg) | 118.7 ± 16.6 | 122.9 ± 16.3 | < 0.001 |
| DBP (mmHg) | 75.1 ± 9.7 | 78.3 ± 11.2 | < 0.001 |
| **Uric acid (mg/dL)** | 4.7 ± 1.0 | 7.5 ± 0.9 | < 0.001 |
| **Fasting blood glucose (mg/dL)** | 101.4 ± 24.7 | 102.7 ± 20.3 | 0.004 |
| **Total cholesterol (mg/dL)** | 191.3 ± 37.1 | 197.4 ± 39.4 | < 0.001 |
| **Triglyceride (mg/dL)** | 126.4 ± 94.5 | 182.5 ± 145.3 | < 0.001 |
| **HDL-C (mg/dL)** | 52.1 ± 12.7 | 46.5 ± 11.1 | < 0.001 |
| **GFR (mL/min/1.73 m<sup>2</sup>)** | 91.5 ± 18.1 | 81.5 ± 19.7 | < 0.001 |
| **Hypertension** |  |  | < 0.001 |
| Normal | 6809 (44.7%) | 684 (28.9%) |  |
| Pre-hypertension | 3696 (24.3%) | 675 (28.5%) |  |
| Hypertension | 4714 (31.0%) | 1006 (42.5%) |  |
| **Diabetes** |  |  | < 0.001 |
| Normal | 9059 (59.5%) | 1158 (49.0%) |  |
| Pre-diabetes | 4198 (27.6%) | 880 (37.2%) |  |
| Diabetes | 1962 (12.9%) | 327 (13.8%) |  |
| **Dyslipidemia** | 6128 (40.3%) | 1389 (58.7%) | < 0.001 |
| **Smoking** |  |  | < 0.001 |
| Non-smoker | 9498 (62.4%) | 1026 (43.4%) |  |
| Ex-smoker | 3524 (23.2%) | 804 (34.0%) |  |
| Current smoker | 2197 (14.4%) | 535 (22.6%) |  |
| **Alcohol drinking** | 6416 (42.2%) | 1327 (56.1%) | < 0.001 |
| **Regular exercise** | 6252 (41.1%) | 1020 (43.1%) | 0.06 |
| **The number of dental caries** |  |  | < 0.001 |
| 0 | 11287 (74.2%) | 1597 (67.5%) |  |
| 1,2 | 2857 (18.8%) | 538 (22.7%) |  |
| ≥3 | 1075 (7.1%) | 230 (9.7%) |  |
| **The number of tooth brushing** |  |  | < 0.001 |
| ≤1 | 1522 (10.0%) | 335 (14.2%) |  |
| 2 | 5682 (37.3%) | 924 (39.1%) |  |
| ≥3 | 8015 (52.7%) | 1106 (46.8%) |  |
| **The use of secondary oral products** | 8441 (55.5%) | 1212 (51.2%) | < 0.001 |
| **Regular dental examination** | 5637 (37.0%) | 788 (33.3%) | 0.001 |

BMI, body mass index; SBP, systolic blood pressure; DBP, diastolic blood pressure; HDL-C,
high-density lipoprotein cholesterol; GFR, glomerular filtration rate;
Data are expressed as mean ± standard deviation or number (%)

### Association between oral health and hyperuricemia

The association between oral health and hyperuricemia is presented in Table 2. The multivariable-adjusted logistic regression analyses were performed. In all subjects, oral health status with dental caries and oral health behaviors including tooth brushing; use of secondary oral products, and regular dental examination, was significantly associated with hyperuricemia (P < 0.05). The adjusted OR and 95% CIs for hyperuricemia comparing more than three dental caries with no dental caries were 1.28 (1.08–1.51). The adjusted OR and 95% CIs for hyperuricemia comparing more than three tooth brushing with under one tooth brushing were 0.78 (0.67–0.91). The adjusted OR and 95 CIs for hyperuricemia in secondary oral products and regular dental examination were 0.91 (0.82–1.00) and 0.86 (0.78–0.95), respectively. When the study was expanded to include sex stratification, the male group exhibited a significant association from oral health and hyperuricemia.

**Table 2.** Odds ratios with 95% confidence intervals for the association between oral health and hyperuricemia.

|  |  | Hyperuricemia<br>(uric acid > 7.0 mg/dL in male, > 6.0 mg/dL in female) |  |  |  |  |  |
| --- | --- | --- | --- | --- | --- | --- | --- |
|  |  | Crude OR | 95% CI | P-value | Adjusted OR* | 95% CI | P-value |
| **All subjects**<br>**(n = 17,584)** |  |  |  |  |  |  |  |
| The number of dental caries | 0 | 1 |  |  | 1 |  |  |
|  | 1,2 | 1.33 | 1.20–1.48 | < 0.001 | 1.14 | 1.02–1.28 | 0.03 |
|  | ≥3 | 1.51 | 1.30–1.76 | < 0.001 | 1.28 | 1.08–1.51 | 0.004 |
| The number of tooth brushing | 1≤ | 1 |  |  | 1 |  |  |
|  | 2 | 0.74 | 0.64–0.85 | < 0.001 | 0.83 | 0.72–0.97 | 0.02 |
|  | ≥3 | 0.63 | 0.55–0.72 | < 0.001 | 0.78 | 0.67–0.91 | 0.001 |
| The use of secondary oral products | No | 1 |  |  | 1 |  |  |
|  | Yes | 0.84 | 0.77–0.92 | < 0.001 | 0.91 | 0.82–1.00 | 0.04 |
| Regular dental examination | No | 1 |  |  | 1 |  |  |
|  | Yes | 0.85 | 0.78–0.93 | < 0.001 | 0.86 | 0.78–0.95 | 0.003 |
| **Male subjects**<br>**(n = 7,831)** |  |  |  |  |  |  |  |
| The number of dental caries | 0 | 1 |  |  | 1 |  |  |
|  | 1,2 | 1.18 | 1.03–1.35 | 0.01 | 1.10 | 0.96–1.27 | 0.17 |
|  | ≥3 | 1.29 | 1.07–1.54 | 0.007 | 1.27 | 1.04–1.55 | 0.02 |
| The number of tooth brushing | 1≤ | 1 |  |  | 1 |  |  |
|  | 2 | 1.02 | 0.87–1.21 | 0.78 | 0.85 | 0.71–1.02 | 0.08 |
|  | ≥3 | 1.03 | 0.87–1.21 | 0.73 | 0.80 | 0.67–0.96 | 0.02 |
| The use of secondary oral products | No | 1 |  |  | 1 |  |  |
|  | Yes | 1.14 | 1.02–1.27 | 0.02 | 0.98 | 0.87–1.11 | 0.77 |
| Regular dental examination | No | 1 |  |  | 1 |  |  |
|  | Yes | 0.92 | 0.82–1.03 | 0.15 | 0.87 | 0.76–0.98 | 0.02 |
| **Female subjects**<br>**(n = 9,753)** |  |  |  |  |  |  |  |
| The number of dental caries | 0 | 1 |  |  | 1 |  |  |
|  | 1,2 | 1.32 | 1.09–1.59 | 0.004 | 1.16 | 0.95–1.42 | 0.15 |
|  | ≥3 | 1.37 | 1.02–1.83 | 0.04 | 1.22 | 0.89–1.67 | 0.23 |
| The number of tooth brushing | 1≤ | 1 |  |  | 1 |  |  |
|  | 2 | 0.55 | 0.43–0.70 | < 0.001 | 0.77 | 0.58–1.02 | 0.07 |
|  | ≥3 | 0.43 | 0.34–0.55 | < 0.001 | 0.80 | 0.60–1.05 | 0.11 |
| The use of secondary oral products | No | 1 |  |  | 1 |  |  |
|  | Yes | 0.67 | 0.58–0.78 | < 0.001 | 0.87 | 0.74–1.03 | 0.10 |
| Regular dental examination | No | 1 |  |  | 1 |  |  |
|  | Yes | 0.76 | 0.65–0.89 | 0.001 | 0.93 | 0.79–1.11 | 0.45 |

OR, odds ratio; CI, confidence interval; BMI, body mass index; GFR, glomerular filtration rate.
\*Logistic model was adjusted for age, sex, income level, BMI, GFR, hypertension, diabetes, dyslipidemia status, smoking status, alcohol consumption, and exercise status.

### Association between the oral health score and hyperuricemia

We calculated the oral health score using oral health status and behaviors. The association between the oral health score and hyperuricemia is presented in Fig 2. There was significant tendency between the oral health score and hyperuricemia regardless of the study population (P for trend < 0.05). Specifically, the adjusted OR and 95% CIs for hyperuricemia comparing score 6 with score 0 were 0.41 (0.27–0.64) in all subjects, 0.56 (0.34–0.93) in male subjects, and 0.23 (0.10–0.50) in female subjects, respectively. In addition, the subgroup analysis based on age is described in Supplementary Table 4. When dividing the study population as median age (52 years), the adjusted OR and 95% CIs for hyperuricemia comparing score 6 with score 0 were 0.61 (0.31–1.21) in under-median age and 0.41 (0.23–0.73) in above-median age, respectively.

**Fig 2.**
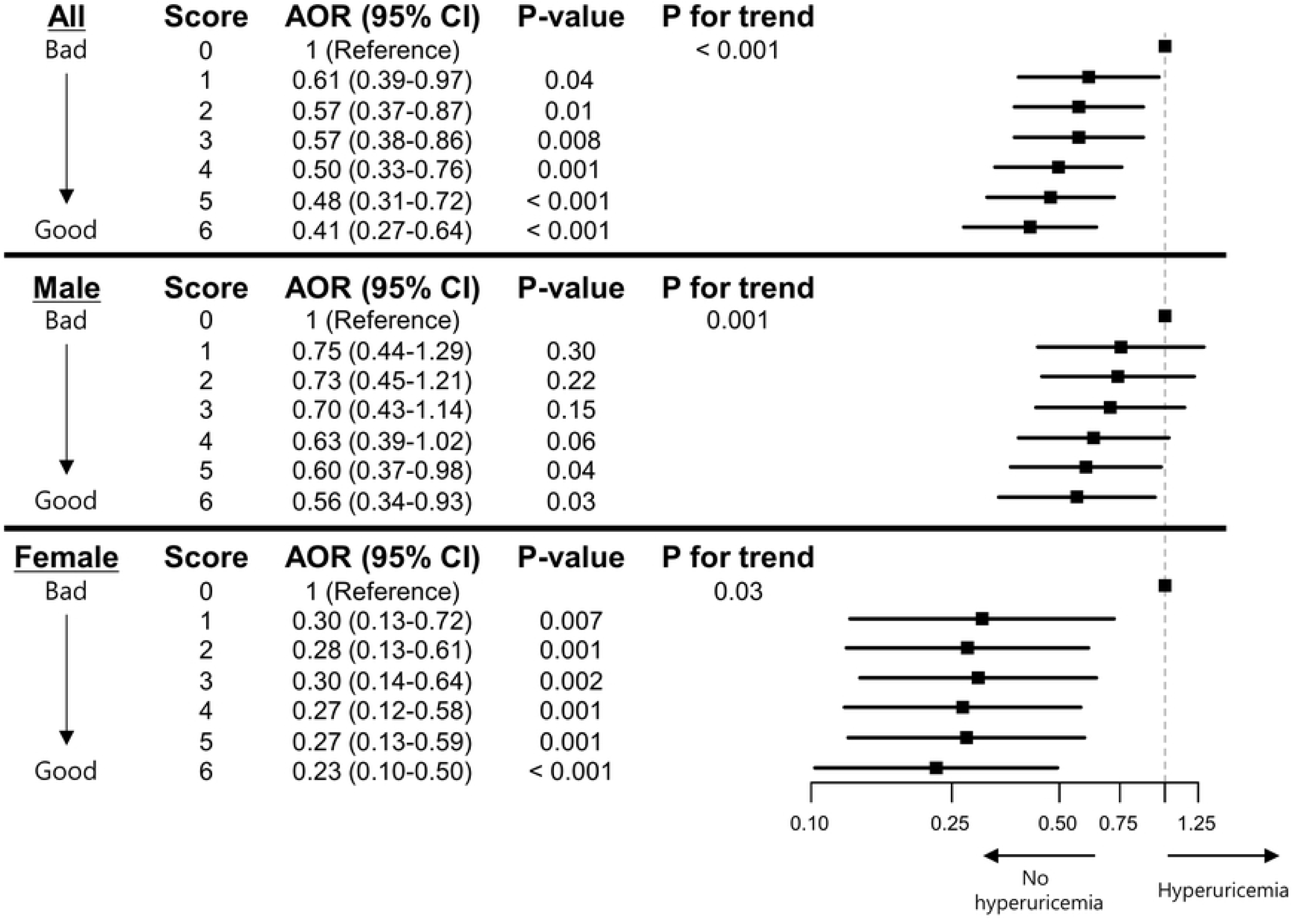
Odds ratios with 95% confidence intervals for the association between oral health score and hyperuricemia. AOR, adjusted odds ratio; CI, confidence interval. *Logistic model was adjusted for age, sex, income level, BMI, GFR, hypertension, diabetes, dyslipidemia status, smoking status, alcohol consumption, and exercise status.

## Discussion

This study investigated the association between oral hygiene and hyperuricemia using a large-scale cross-sectional study with KNHANES data. The number of dental caries was associated with hyperuricemia and oral hygiene behaviors, such as regular tooth brushing; use of secondary oral products; and regular dental examination, were also related with hyperuricemia in all subjects (Table 2). Furthermore, we applied the oral health score by grading oral health-related variables and summing them altogether. As a result, the significant association between the oral health score and hyperuricemia was also confirmed (Fig 2). In gender-based subgroup analysis, we could notice a tendency that the higher oral health score was associated with the lower incidence of hyperuricemia regardless of sex. In age-based subgroup analysis, a similar correlation between the oral health score and hyperuricemia was maintained in both groups, but it was statistically valid only in the elderly group (S4 Table).

The association between oral disease and hyperuricemia has remained controversial; although, there have been several previous studies about them. Recently, Byun et al. analyzed the relationship between periodontitis and hyperuricemia in a large population and reported that the two diseases were not significantly correlated [19]. On the other hand, Banu et al. reported that blood uric acid levels in patients with periodontitis were higher than those in normal subjects [20]. The former study may have had data reliability issues because it used data acquired over an extended period (from 2004 to 2016). In case of the latter study, there may have been problems of including a limited number of patients as samples. In this study, we used data from the KNHANES 2016–2019 database with the same protocol within a relatively smaller period and secured a sufficient number of subjects for statistical analysis, which could be a strength of this study.

Although a clear mechanism between oral hygiene and hyperuricemia is unknown, several perspectives can be derived from previous studies. First, the change in oral microbiota caused by poor oral hygiene can be mentioned, as oral microbiota is known for its association with various diseases [21]. In a previous study, Prevotella Intermedia, which is known as one of the causative bacteria of tooth decay and periodontitis, was abundant in oral microbiota of gout patients [22]. In addition, the color of the root of the tongue, which may be related to oral health and oral microbiota, is indicative of hyperuricemia [23]. Furthermore, it has been reported that abnormal findings or oral microbiota are also associated with severe asthma, cardiovascular disease, diabetes, and rheumatoid arthritis [24–30]. Poor oral hygiene may cause changes in oral microbiota, which may explain the association between oral hygiene and hyperuricemia [31]. Second, oral disease itself is known to have the association with metabolic disorders. Consistent with the results in this study, the previous meta-analysis study also exhibited the association between periodontitis and hypertension [32]. In addition, Kobayashi et al. reported that oral hygiene is associated with metabolic syndrome [33]. According to a recent randomized controlled trial, the risk of metabolic syndrome was lower in the group with oral hygiene intervention compared to the group without intervention [34]. Taken all together, it is possible to infer that oral hygiene may be related to hyperuricemia with metabolic disorders.

This study has some limitations. First, the oral health-related variables do not include the presence of periodontitis, which is one of the common clinical manifestations of an individual’s oral hygiene. However, in the case of periodontitis, dental examination and radiological examination are necessary. Furthermore, since the severity of periodontitis may vary from patient to patient, we quantitatively evaluated the oral health condition by counting the number of dental caries, instead. Second, oral health status and behaviors were investigated using a self-reporting questionnaire. Third, there is a possibility of unadjusted covariates that are not analyzed in this study. Finally, this cross-sectional study was to infer the association between oral hygiene and hyperuricemia, but it could not establish the cause-and-effect relationship. We expect that a future study will be able to identify the predecessor relationship between oral hygiene and hyperuricemia.

## Conclusion

In this study, we demonstrated that oral health status and behaviors are associated with hyperuricemia, especially in males. In addition, when subjects are grouped by the oral health scoring system, the prevalence of hyperuricemia is lower in groups with a better oral health score. Additional complementary studies are needed to confirm the definite association between oral health and hyperuricemia.

## Data Availability

Data are available from the Korea National Health and Nutrition Examination Survey (KNHANES), performed by the Korea Centers for Disease Control and Prevention (KCDCP), and are freely available from the website (https://knhanes.kdca.go.kr/knhanes/eng/index.do).

## Supporting information

**S1 Table. The schematic table for oral health score**.

**S2 Table. Baseline characteristics of male subjects according to uric acid level**.

BMI, body mass index; SBP, systolic blood pressure; DBP, diastolic blood pressure; HDL-C, high-density lipoprotein cholesterol; GFR, glomerular filtration rate;

Data are expressed as mean ± standard deviation or number (%).

**S3 Table. Baseline characteristics of female subjects according to uric acid level**.

BMI, body mass index; SBP, systolic blood pressure; DBP, diastolic blood pressure; HDL-C, high-density lipoprotein cholesterol; GFR, glomerular filtration rate;

Data are expressed as mean ± standard deviation or number (%).

**S4 Table. Subgroup analysis based on median age (52 years) for the association between oral health score and hyperuricemia**.

OR, odds ratio; CI, confidence interval; BMI, body mass index; GFR, glomerular filtration rate. *Logistic model was adjusted for age, sex, income level, BMI, GFR, hypertension, diabetes, dyslipidemia status, smoking status, alcohol consumption, and exercise status.

